# Identification and characterization of collagen XXIII alpha 1 as a novel risk factor for eczema herpeticum

**DOI:** 10.1101/2024.07.13.24310236

**Authors:** Shruti Chopra, Lennart M. Roesner, Katinka Döhner, Jana Zeitvogel, Stephan Traidl, Elke Rodriguez, Inken Harder, Wolfgang Lieb, Stephan Weidinger, Thomas F. Schulz, Beate Sodeik, Thomas Werfel

## Abstract

Eczema herpeticum (EH) is a potentially life-threatening disseminated skin infection caused by herpes simplex virus (HSV) in a subset of patients with atopic dermatitis (AD). Genetic factors play a pivotal role in EH development. Herein, we identify a single nucleotide polymorphism (SNP) rs2973744 in the gene encoding collagen XXIII alpha 1 chain (Col23a1) as a novel genetic risk factor for EH using whole exome sequencing. EH-patient-derived primary keratinocytes carrying the SNP rs2973744 show elevated *COL23A1* mRNA and total protein levels as well as increased susceptibility to HSV-1. We discover that increasing Col23a1 levels enhances HSV-1 infection in human keratinocytes. The transcriptomic analysis unveils that *COL23A1* overexpression dampens keratinocyte immune responses, thereby elucidating the molecular mechanism underlying exacerbated HSV-1 susceptibility. Our findings report a novel potential screening marker and therapeutic target for EH and reveal Col23a1’s unexplored role in HSV-1 pathogenesis.

## Introduction

Atopic dermatitis (AD) is a common chronic inflammatory skin disease that affects 20% of children and 2-5% of adults worldwide ^1, 2, 3^. It is a multi-factorial disease influenced by genetic, immunological and environmental factors. Patients suffering from AD have an impaired skin barrier with type 2-dominated immune responses and heightened allergen sensitization. This presumably renders them more susceptible to bacterial and viral infections, including infections with herpes simplex virus (HSV) ^4, 5, 6, 7^. Around 67% and 13% of humans worldwide have been exposed to HSV-1 and HSV-2 respectively, however, most of them develop no or only minor symptoms ^8^.

HSV-1 is an enveloped double-stranded DNA virus that infects stratified epithelial cells of skin, mucosa or cornea, where the predominant cell type is keratinocytes. HSV-1 enters keratinocytes either by fusion at the cell membrane or through the endocytic pathway ^9, 10^. After fusion of the viral envelope with a cellular membrane, incoming capsids are transported to the nucleus where viral transcription takes place in a regulated cascade-like manner, starting with the immediate-early genes, followed by early and finally late genes ^11, 12, 13, 14^. After primary infection of stratified epithelial cells, HSV-1 spreads to neurons and establishes a lifelong latency in trigeminal or dorsal root ganglia. Upon reactivation, it can be transported back to the original dermatome in the skin or mucosa ^11, 13, 15, 16, 17^.

In a subgroup of AD patients, HSV can cause severe disseminated infections, referred to as eczema herpeticum (EH) ^7, 18^. EH can lead to life-threatening encephalitis or even death if left untreated or misdiagnosed, therefore, EH is considered as a dermatological emergency ^19^. Approximately 22% of moderately to severely affected AD patients report at least one episode of EH ^7, 18, 20^. Notably, over 50% of these individuals have had recurrent episodes. These findings suggest genetic predispositions as contributing factors in EH development.

Previous studies identified several genetic factors associated with EH ^21^. Particularly, the R501X mutation in filaggrin, an important structural protein involved in skin barrier function, increased the risk of developing EH threefold in AD patients ^22, 23^. SNPs in CLDN1 and CLDN16 encoding the tight junction proteins claudin-1 and claudin-16, and in SIDT2 and RBBP8NL encoding SID1 transmembrane family member 2 and RBBP8 N-terminal like, are also associated with an increased risk of EH in AD patients, and silencing CLDN1, SIDT2 or RBBP8NL increases HSV-1 infection ^24, 25, 26^.

Here, we performed whole exome sequencing in AD patients without (ADEH-) or with a history of EH (ADEH+) to discover further genetic variants associated with EH. Our study identified a novel heterozygous SNP in *COL23A1*, rs2973744, to be significantly associated with EH development (χ^2^-test: *p=0.034*). This SNP rs2973744 in *COL23A1* causes a thymine to cytosine substitution at a splice donor site of the transcript allele between exons 10 and 11 leading to enhanced protein expression in keratinocytes. Moreover, we report that increased expression of Col23a1 enhances HSV-1 infection in keratinocytes by downregulating genes involved in immune responses.

## Results

### Whole exome sequencing revealed a novel single nucleotide polymorphism (SNP) in *COL23A1* rs2973744 associated with eczema herpeticum

To identify novel genetic risk factors for EH, we performed next-generation whole exome sequencing (WES) of nine ADEH+ patients and their respective first-degree relatives and identified the single-nucleotide polymorphism (SNP) rs2973744 in the gene *COL23A1* located on chromosome 5q35. Three out of the nine ADEH+ patients and one out of the unaffected healthy relatives were carriers of the heterozygous SNP rs2973744. This genetic variant was further validated in a larger cohort comprising 117 ADEH+ and 117 ADEH- patients ^27^ (GENEVA cohort and RESIST HSV/AD cohort), and 118 healthy controls ^28^ (POPGEN cohort). DNA was obtained from each group and Taqman PCR was used to investigate the occurrence of this variant. As shown in Table 1, the occurrence of the SNP rs2973744 was observed in 5% of ADEH+ patients, 1.6% of healthy donors and 0% of ADEH- patients, thus making rs2973744 significantly associated with EH in this group of AD patients (ꭓ^2^ test: *p=0.034*).

**Table 1.** Frequency of the heterozygous SNP rs2973744.

|  |  |
| --- | --- |
| ADEH+ (n = 117) | 5.0% |
| ADEH- (n = 117) | 0.0% |
| Healthy (n = 118) | 1.6% |
| Chi-square, df | 6,762.2 |
| P value | 0.034 |
| P value summary | * |

### Primary human keratinocytes from an ADEH+ patient with SNP rs2973744 expressed higher Col23a1 levels and were more susceptible to HSV-1 than keratinocytes from ADEH- patients

Since epidermal keratinocytes are the major entry portal for HSV-1 infections of the skin ^14^, we investigated the role of Col23a1 in HSV-1 infections of keratinocytes. Therefore, we first analyzed the susceptibility of primary keratinocytes isolated from hair follicles of ADEH- or ADEH+ patients to a recombinant HSV-1 strain expressing green fluorescent protein (GFP) as a reporter [HSV1-GFP, multiplicity of infection (MOI) of 0.75 plaque-forming units (PFU)/cell]. To avoid the effect of differentiation stages on HSV-1 susceptibility between primary hair keratinocytes derived from different patients, cells were pre-stimulated with 1.4 mM CaCl_2_ for 24 h before each experimental set-up. HSV1-GFP expression was higher in keratinocytes from an ADEH+ patient with the *COL23A1* SNP than in keratinocytes derived from a healthy donor, or ADEH- patients without or with the SNP (Figure 1a). To investigate whether *COL23A1* expression differed between the groups and could possibly be a factor affecting HSV-1 susceptibility, we measured their relative *COL23A1* transcript and total protein levels. Primary keratinocytes isolated from the ADEH+ patient carrying the SNP rs2973744 had the highest *COL23A1* mRNA and protein levels as confirmed by RT-qPCR and Western blotting respectively. (Figure 1b-d). In keratinocytes carrying the SNP the 75 kDa band was stronger than the 60 kDa band, while in healthy keratinocytes the 60 kDa band was more prominent than the 75 kDa band. Moreover, keratinocytes harbouring the SNP had an additional band between 60 and 75 kDa, which was not observed in the keratinocytes without the SNP. These data suggest that the SNP at the splice donor site results in differentially processed *COL23A1* mRNA forms, possibly due to alternative splicing. In summary, increased total Col23a1 levels in keratinocytes from an ADEH+ donor with the SNP rs2973744 correlated with more efficient HSV-1 infection suggesting that this SNP together with other genetic factors associated with EH increases *COL23A1* expression and susceptibility to HSV-1.

**Figure 1.**
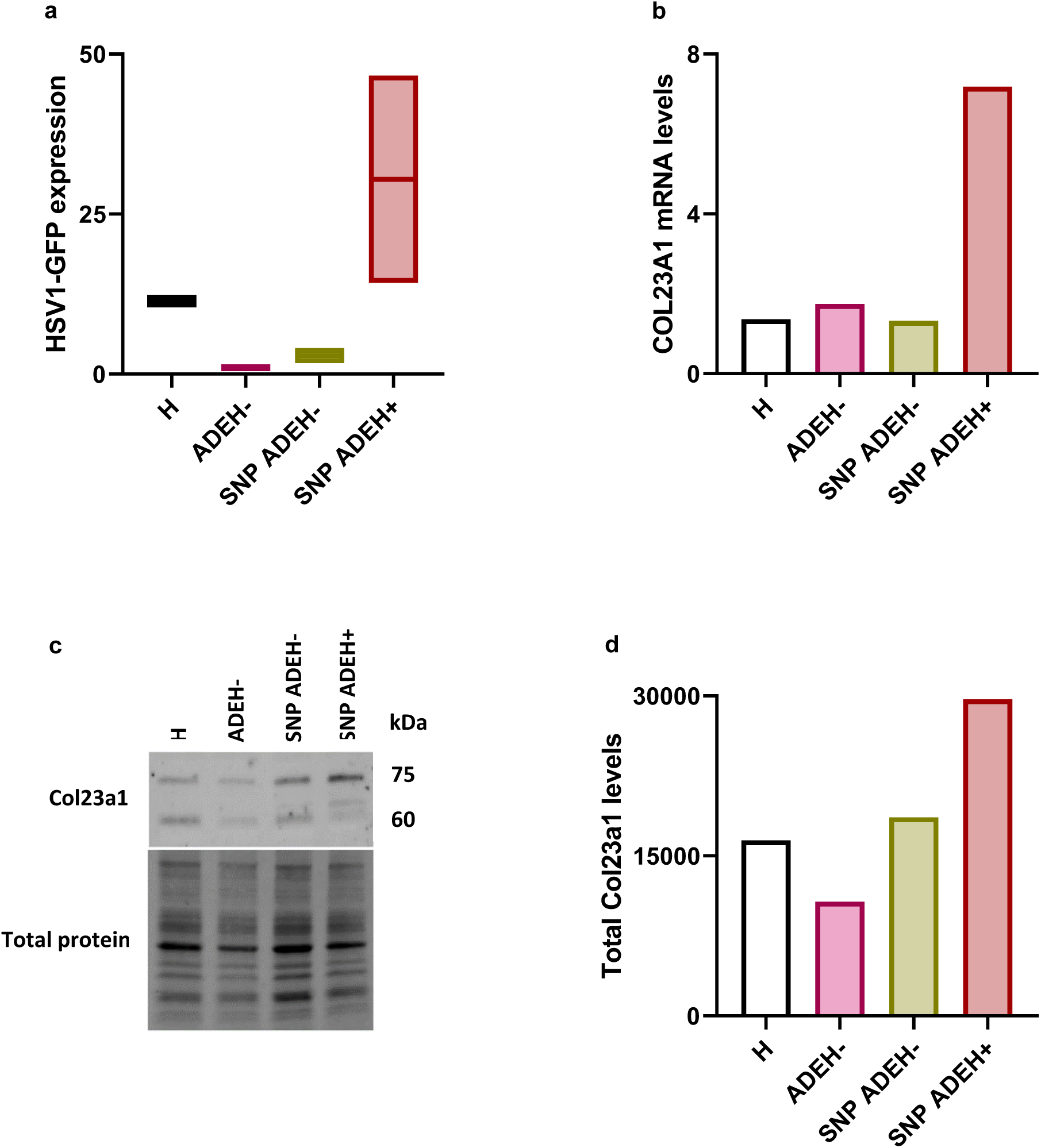
Primary keratinocytes derived from an ADEH+ patient are more susceptible to HSV-1 and express more *COL23A1* mRNA and total protein levels than those from ADEH- patients. **a** Primary hair keratinocytes isolated from a healthy control (H; n = 1), an AD patient without a history of EH without (ADEH-; n = 1) and with the rs2973744 SNP (SNP ADEH-; n = 1) and an AD patient with a history of EH carrying the rs2973744 SNP (SNP ADEH+; n = 1) were infected with 0.75 pfu/cell of HSV1-GFP. 20 h post infection HSV1-GFP expression was measured by flow cytometry. Data were normalized to the mean of infected ADEH- cells (n = 2 independently repeated experiments, Box & whiskers represent 10-90 percentile). **b** *COL23A1* mRNA levels in primary hair keratinocytes from (1A) were quantified by RT-qPCR and normalized to the housekeeping gene *RPS20* (Interleaved bars; n = 1). **c** Total Col23a1 protein levels in primary hair keratinocytes from **a** were visualized using Western blot in comparison to total protein. **d** Col23a1 protein levels visualized in **c** were quantified and normalized to total protein levels (Interleaved bars; n = 1).

### Inhibiting furin enhanced Col23a1 levels on the surface of primary keratinocytes and their susceptibility to HSV-1

Collagen XXIII alpha 1 chain exists in a full-length membrane-bound form and a short soluble ectodomain. The cleavage of Col23a1 in the Golgi/*trans*-Golgi network is catalyzed by proteases, primarily by the enzyme furin ^29^. Since most of the Col23a1 is shed in cultured cells ^30^, we blocked its shedding and analyzed by flow cytometry how enhanced cell surface levels of Col23a1 affect HSV-1 infection. We first treated keratinocytes with 75 µM furin inhibitor I (FI) for 20 h and then infected them with 0.75 pfu/cell of HSV1- GFP for 20 h in the absence of this inhibitor. FI significantly increased cell surface levels of Col23a1 (Figure 2a) and HSV1-GFP expression in primary keratinocytes (Figure 2b) by 2 and 2.5-fold, respectively, indicating that elevated Col23a1 levels on the cell surface might promote HSV-1 infection.

**Figure 2.**
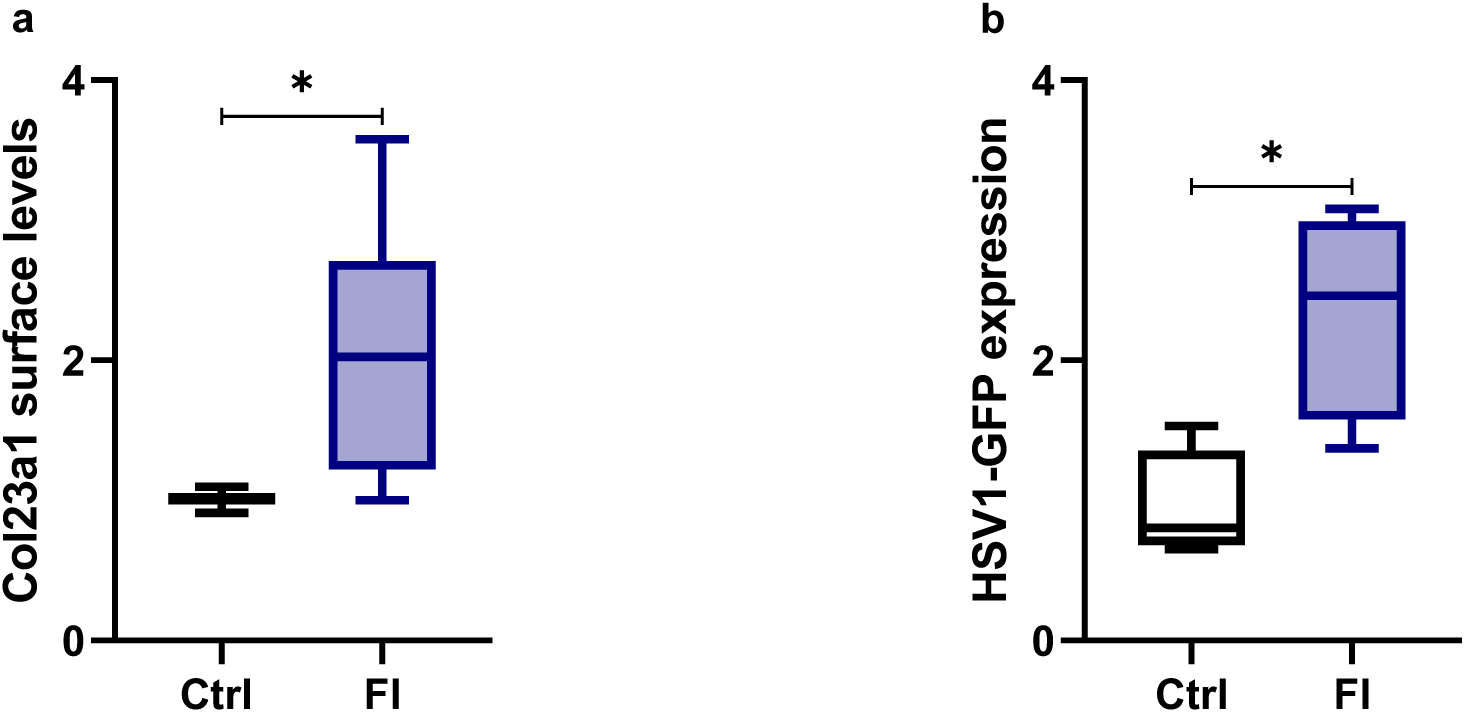
Furin inhibition increases Col23a1 levels on the surface of keratinocytes and enhances their susceptibility to HSV-1. **a** Primary foreskin keratinocytes were stained for Col23a1 after 20 h in the absence (Ctrl) or presence (FI) of furin inhibitor I (FI, 75 µM) and Col23a1 surface levels were analyzed by flow cytometry (n = 6 different donors). Statistical significance was analyzed with Paired t-test, t(5) = 2.933. **b** Primary foreskin keratinocytes were pre-incubated with FI as described in **a** and then infected with 0.75 pfu/cell of HSV1-GFP for 20 h in the absence of FI. The effect of FI treatment on HSV1-GFP expression was measured by flow cytometry. Data was normalized to mean of infected Ctrl cells (Box & whiskers represent 10-90 percentile; n = 4 different donors). Statistical significance was analyzed with Paired t-test, t(3) = 5.680 ; \**P* < 0.05; ns, not significant.

### Stable overexpression of *COL23A1* in HaCaT cells enhanced susceptibility to HSV-1

To confirm the effect of higher Col23a1 expression on enhanced susceptibility to HSV infection we developed a cell culture model of stable *COL23A1* overexpression in the human immortal keratinocyte cell line HaCaT. After transduction with control (ctrl) or *COL23A1* encoding lentiviral vectors, HaCaT cells overexpressed *COL23A1* at the mRNA (Figure 3a) and protein level (Figure 3b, 3c) as revealed by RT-qPCR, flow cytometry and Western blot, respectively. To discern the effect of Col23a1 on HSV-1 infection, we infected ctrl or *COL23A1* overexpressing HaCaT cells with 1 pfu/cell of a recombinant HSV-1 strain expressing mCherry (Che) as a reporter (HSV1-Che). The susceptibility to HSV-1 was measured by quantifying mCherry expression at 20 hpi with flow cytometry. *COL23A1* overexpression significantly increased HSV1-Che expression by about 1.6-fold (Figure 3d), which is consistent with our FI data.

**Figure 3.**
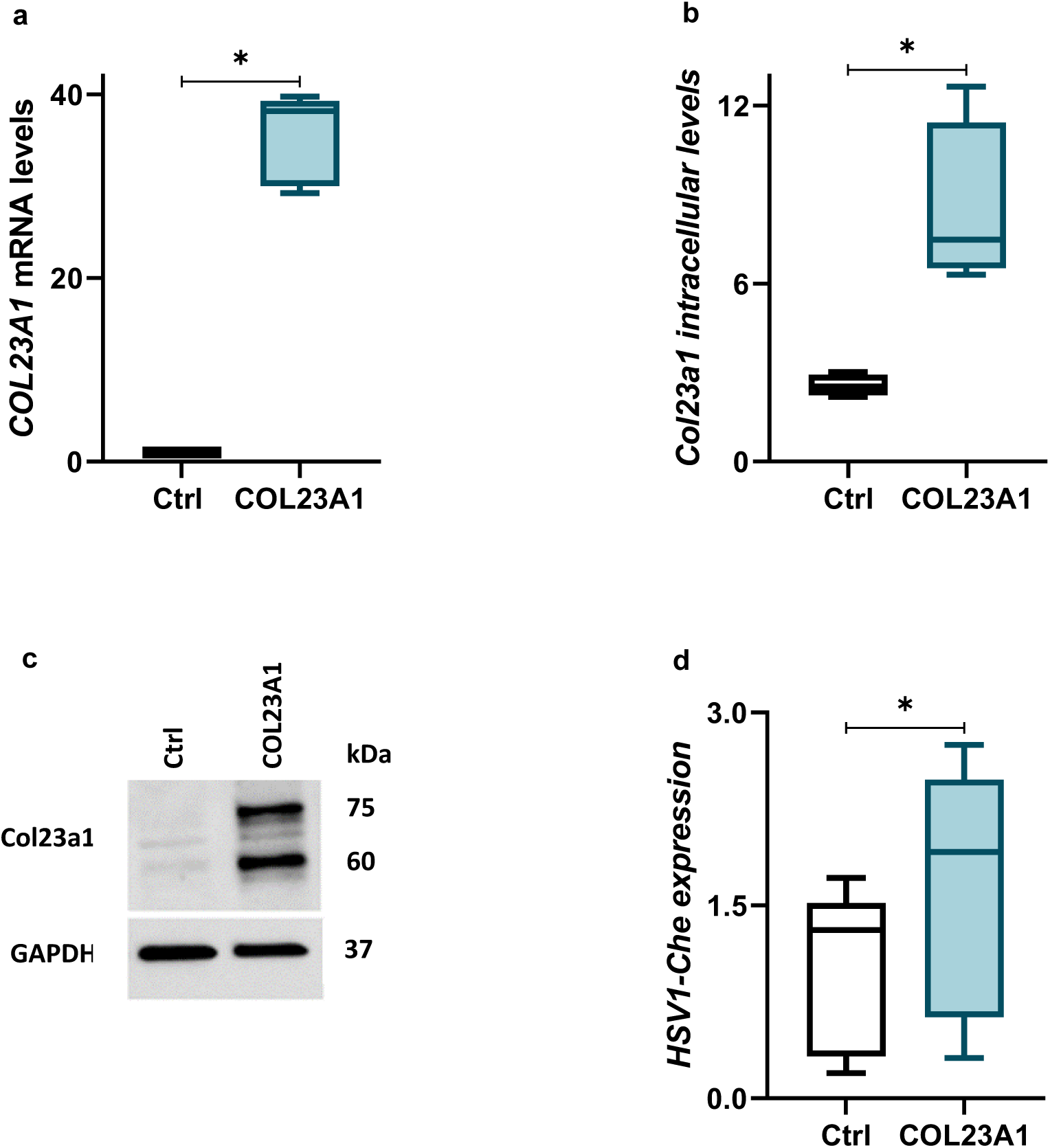
HaCaT cells are more susceptible to HSV-1 after *COL23A1* overexpression. *COL23A1* overexpression was confirmed by quantifying **a** *COL23A1* mRNA levels by RT-qPCR and normalizing to the housekeeping gene *RPS20* (n = 6 independently repeated experiments); Statistical significance was analyzed with One sample Wilcoxon test, **b** intracellular Col23a1 levels by flow cytometry and normalizing to isotype control (n = 4 independently repeated experiments); Statistical significance was analyzed with Paired t-test, t(3) = 4.089 and **c** total Col23a1 protein levels using Western blot in comparison to the housekeeping protein GAPDH (n = 4 independently repeated experiments). **d** Control and *COL23A1* overexpressing HaCaT cells were infected with 1 pfu/cell of HSV1-Che. At 20 hpi, mCherry expression was measured by flow cytometry and normalized to the mean of infected control cells (n = 5 independently repeated experiments; Box & whiskers represent 10-90 percentile). Statistical significance was analyzed with Paired t-test, t(4) = 3.894. \**P* < 0.05; \*\**P* < 0.01. Ctrl= HaCaT cells stably transduced with control lentiviral particles; COL23A1= HaCaT cells stably transduced with *COL23A1* lentiviral particles.

### HSV-1 cell-to-cell spread increased upon *COL23A1* overexpression in HaCaT cells

After 20 h of infection with an MOI of 1 pfu/cell, the virus released from the primarily infected cells has presumably spread to neighboring cells resulting in a second, if not a third, infection cycle. To investigate whether the higher susceptibility of *COL23A1* overexpressing cells was due to increased cell-to-cell spread we infected the cells at very low MOIs of HSV1-Che and quantified the number, size, and intensity of plaques. Interestingly, plaque numbers were increased approximately 2-fold by trend and plaque size and mCherry intensity were slightly but significantly increased in *COL23A1* overexpressing HaCaT cells (Figure 4a-d) thereby indicating that Col23a1 enhanced both initial HSV-1 infection of HaCaT cells as well as cell-to-cell spread.

**Figure 4.**
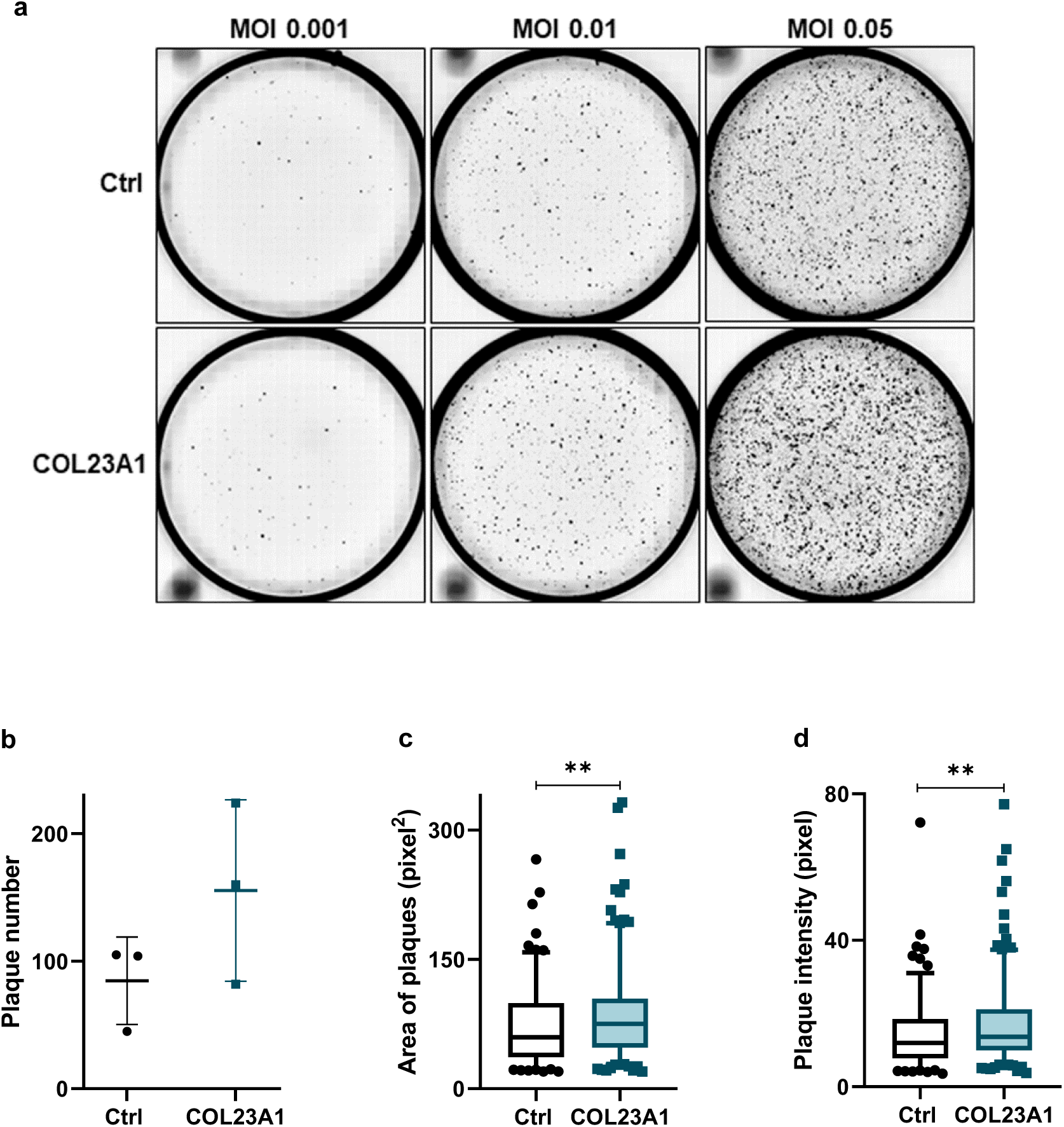
*COL23A1* overexpression increases HSV-1 cell-to-cell spread in HaCaT cells. **a** Control and *COL23A1* overexpressing cells were infected with 0.001, 0.01 or 0.05 pfu/cell of HSV1-Che. mCherry signals were visualized by automated microscopy (n = 3 independently repeated experiments). **b-d** Quantitation of plaque number (**b**, scatter dot plot, mean ± standard deviation [SD]), area and intensity (**c- d**, Box & whiskers represent 10-90 percentile; n = 45-224 plaques at MOI 0.01) were carried out using Cell Profiler. Statistical significances were analyzed with Mann-Whitney test; \*\**P* < 0.01. Ctrl= HaCaT cells stably transduced with control lentiviral particles; COL23A1= HaCaT cells stably transduced with *COL23A1* lentiviral particles.

### *COL23A1* overexpression enhanced HSV-1 gene expression in HaCaT cells

After injection of the HSV-1 genome into the nucleus of an infected cell, it is transcribed in a cascade-like manner starting with immediate-early genes (IE), followed by early (E) and late genes (L) ^12^. Products of IE genes are required for the transcription of E genes, which then encode proteins required for viral DNA synthesis. The products encoded by IE and E genes are further employed for efficient L gene transcription. To better understand which stage of HSV-1 gene expression is affected upon *COL23A1* overexpression, we infected control and *COL23A1* overexpressing HaCaT cells with MOI 5 of HSV-1 and harvested at different time points (2 hpi, 4 hpi, 6 hpi and 8 hpi). We then quantified mRNA expression of two late (*UL19, UL27*), early (*UL23, UL42*) and immediate-early (*UL54, US12*) genes by RT-qPCR (Supplementary Figure S1). We selected 6 hpi and 8 hpi time-points to analyze HSV-1 gene expression in three independent experiments (Figure 5a-f). Interestingly, HSV-1 genes of all the three kinetic classes were upregulated upon *COL23A1* overexpression, thus indicating that *COL23A1* overexpression increases HSV-1 infection at or upstream of immediate-early gene transcription.

**Figure 5.**
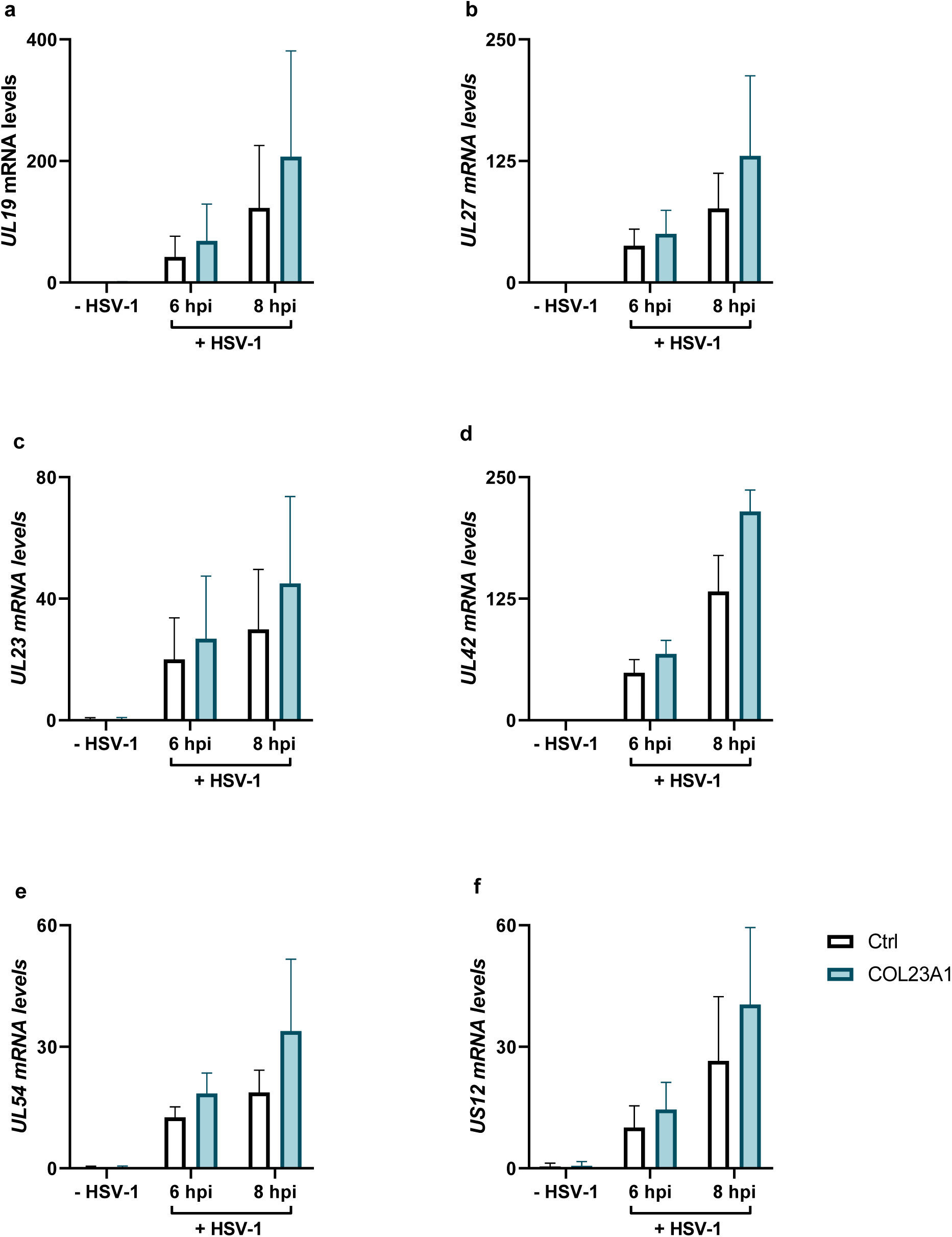
HSV-1 gene expression is more efficient in *COL23A1* overexpressing HaCaT cells. **a-f** Control and *COL23A1* overexpressing HaCaT cells were infected with 5 pfu/cell of HSV-1. *UL19, UL27, UL23, UL42, UL54 and US12* mRNA levels were measured at 6 hpi and 8 hpi by RT-qPCR and normalized to the housekeeping gene *RPS20* (black bars, mean ± standard deviation [SD]; n = 3 independently repeated experiments). Ctrl= HaCaT cells stably transduced with control lentiviral particles; COL23A1= HaCaT cells stably transduced with *COL23A1* lentiviral particles.

### *COL23A1* overexpression in HaCaT cells dampened the expression of genes involved in immune responses

To understand the underlying mechanism by which *COL23A1* overexpression increases HSV-1 susceptibility, cell-to-cell spread and viral gene expression, we analyzed the global transcriptomics of these cells using bulk RNA-seq analysis. We infected control and *COL23A1* overexpressing HaCaT cells with MOI 5 of HSV-1 and isolated their RNA after 2 hpi and 6 hpi. After generating and sequencing cDNA libraries we detected 225 differentially regulated genes in pooled HSV-1 infected and uninfected *COL23A1* overexpressing HaCaT cells as compared to control cells (*p*-adj*≤0.05*). Out of those 225 genes, we shortlisted 189 genes with log 2-fold change [log2(FC)] value of >0.5 or <-0.5 as shown in the volcano plot (Figure 6a). As expected, the highest change was observed in *COL23A1* expression with a value of 8.04 log2(FC) upon *COL23A1* overexpression (not shown). Interestingly, several genes involved in innate or adaptive immune responses like *IL1R1*, *IL32*, *TLR4*, *CFH*, *C3*, *S100A9*, *IRF1*, *ADAM23* were significantly downregulated and *TNC* and *SPINK5*, two genes associated with AD ^31, 32, 33, 34^, were significantly upregulated (Figure 6b). Kyoto Encyclopedia of Genes and Genomes (KEGG) and Gene ontology (GO) pathway analyses reflected a significant suppression of immune responses, inflammatory responses and their regulation, cytokine production as well as regulation of cell adhesion upon *COL23A1* upregulation (Figure 6c-d).

**Figure 6.**
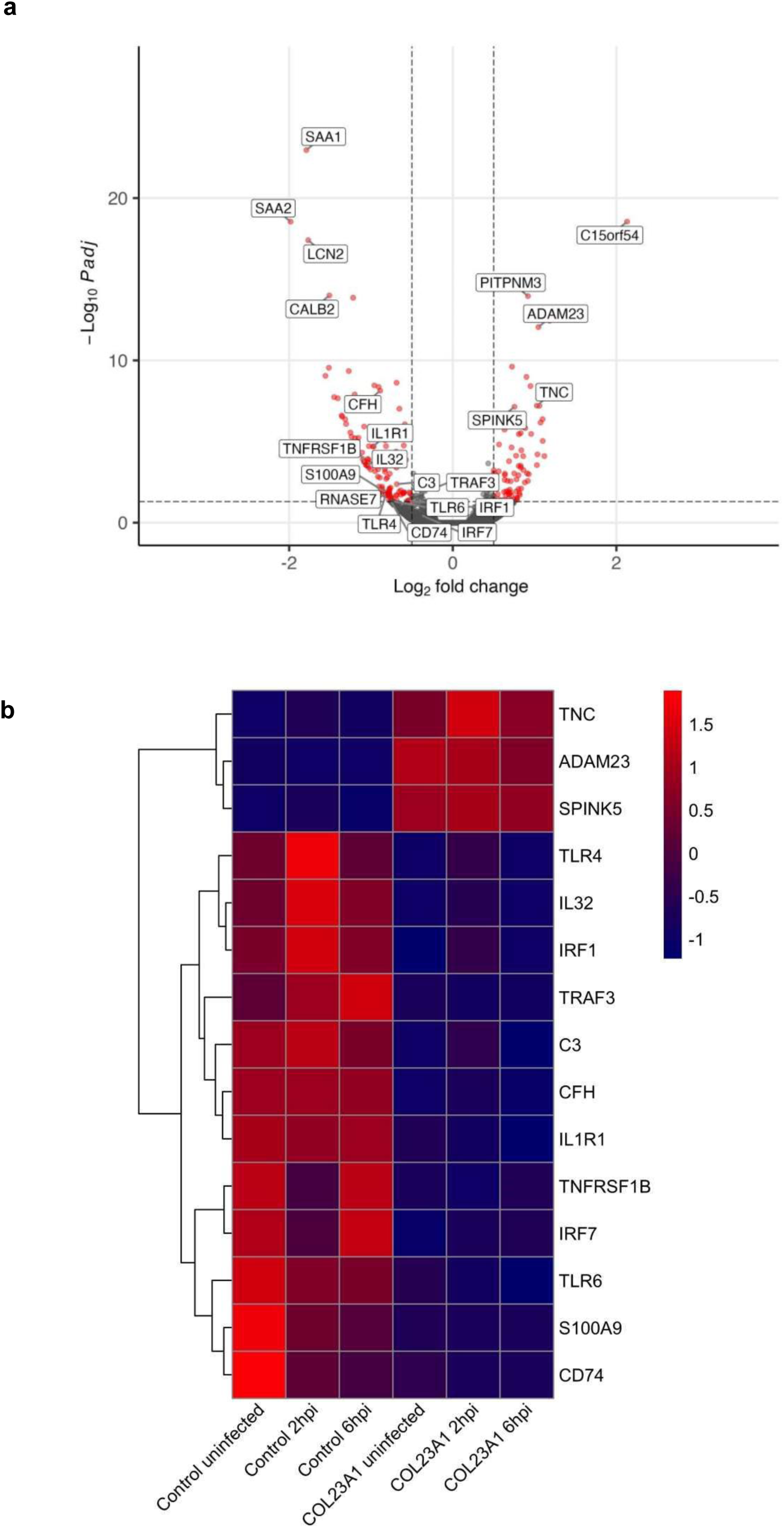
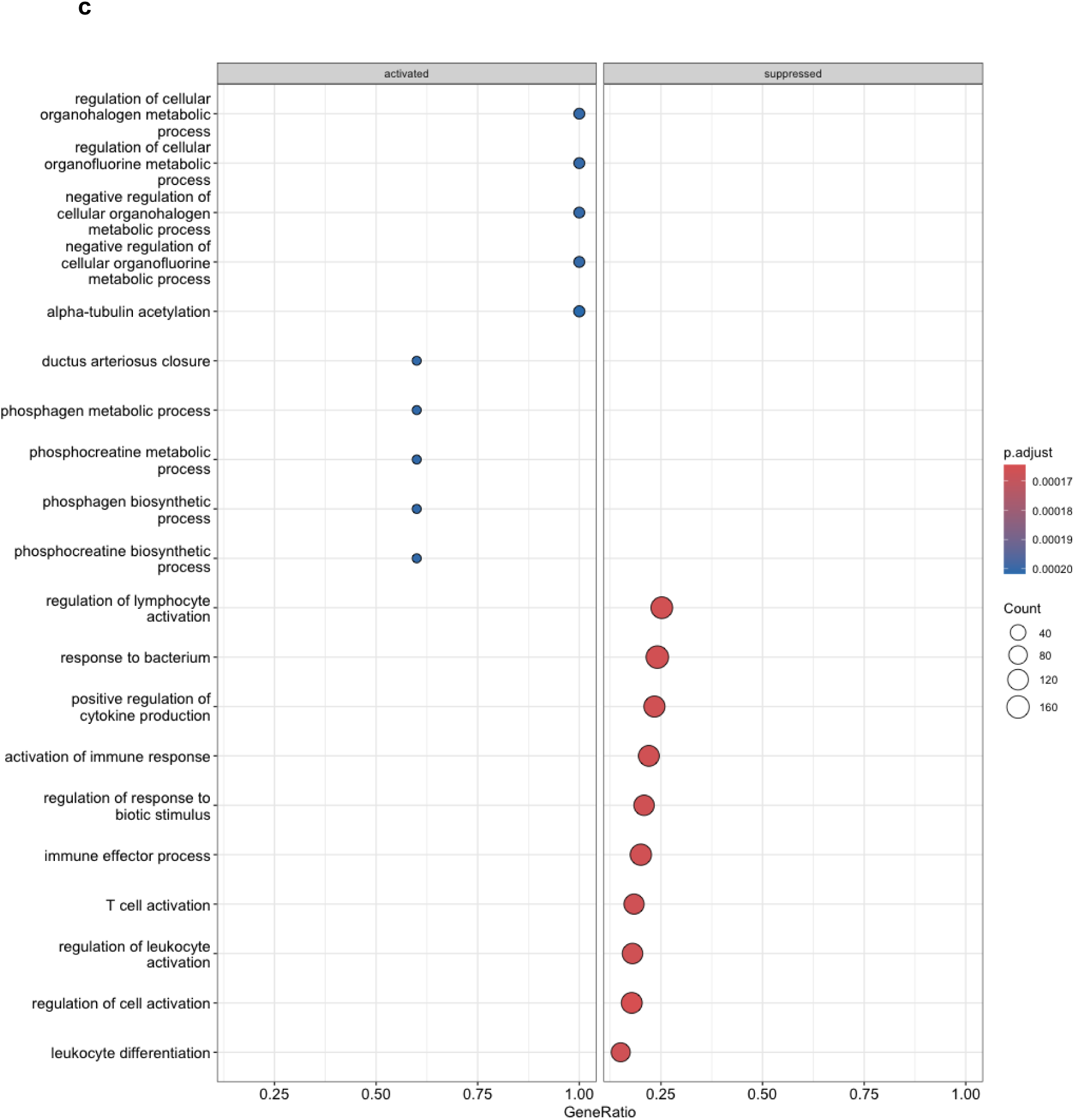
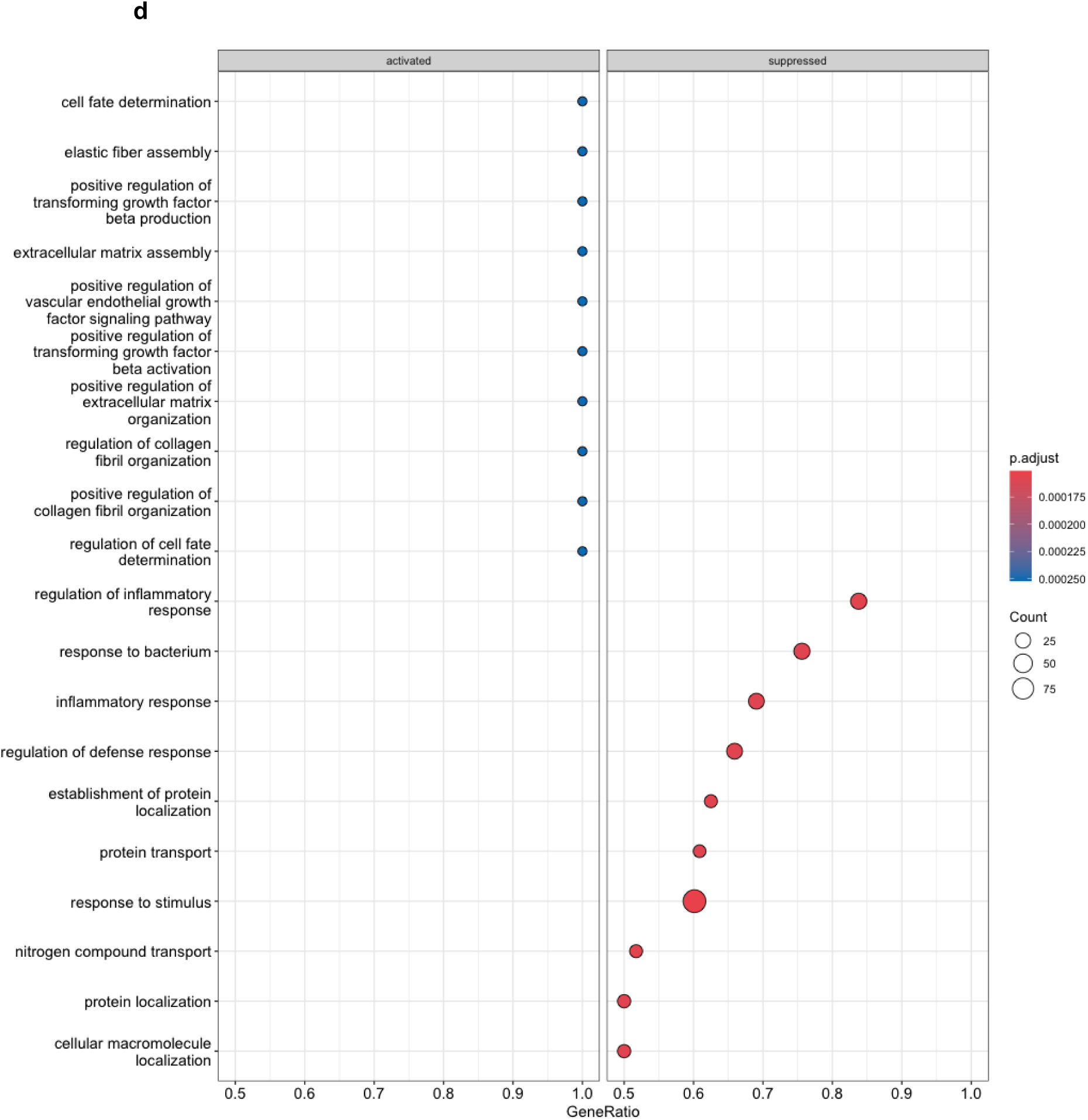
Global transcriptomic changes after *COL23A1* overexpression in HaCaT cells using bulk RNA-seq. HaCaT Control and *COL23A1* overexpressing cells were infected with 5 pfu/cell of HSV-1 and RNA was isolated at 2 hpi and 6 hpi. **a** 189 differentially expressed genes with log2(FC) of >0.5 or <-0.5 were visualized using a volcano plot. **b** Heatmap illustrated the differential expression of selected genes involved in immune response or AD in control and *COL23A1* overexpressing HaCaT cells. Bubble plots depicting significantly enriched KEGG (**c**) and GO pathways (**d**) after *COL23A1* overexpression were generated by using R package clusterProfiler.

RT-qPCR confirmed the downregulation of several host genes involved in an effective immune response and upregulation of those associated with AD in uninfected *COL23A1* overexpressing cells as compared to control cells (Figure 7), suggesting that *COL23A1* overexpression enhances the susceptibility of keratinocytes to HSV-1 by dampening immune responses.

**Figure 7.**
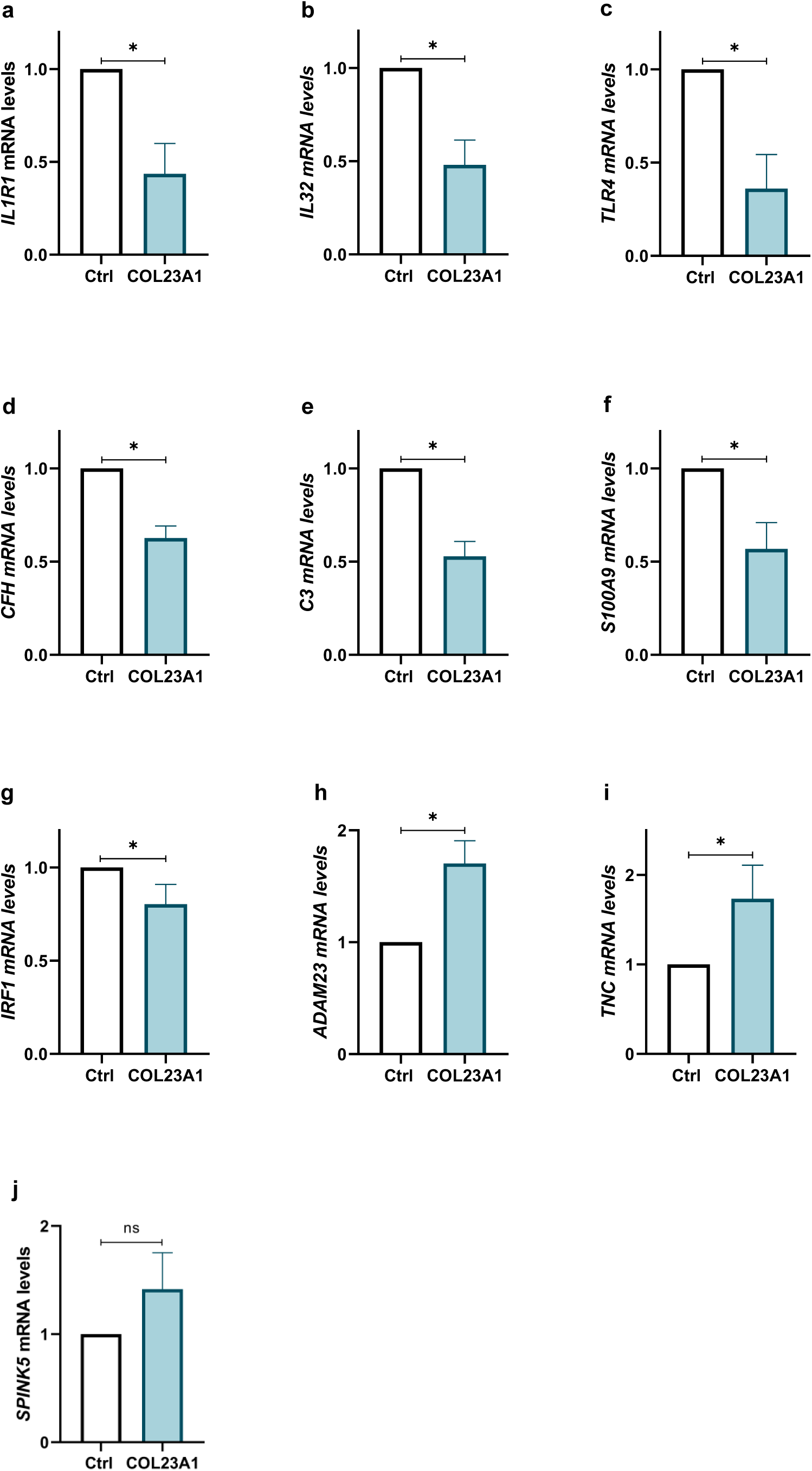
Confirmation of hits revealed in bulk RNA-seq data using RT-qPCR. **a-f** *IL1R1*, *IL32*, *TLR4*, *CFH*, *C3*, *S100A9*, *IRF1*, *ADAM23*, *TNC*, and *SPINK5* mRNA levels were measured in uninfected control and *COL23A1* overexpressing cells by RT-qPCR and normalized to the housekeeping gene *RPS20* (black bars, mean ± standard deviation [SD]; n = 6 independently repeated experiments). Statistical significances were analyzed with One sample Wilcoxon test; \**P* < 0.05; ns, not significant. Ctrl= HaCaT cells stably transduced with control lentiviral particles; COL23A1= HaCaT cells stably transduced with *COL23A1* lentiviral particles.

## Discussion

Previous studies identified various genetic risk factors involved in EH using GeneChip profiling, genotyping, targeted resequencing approaches and WGS ^22, 24, 35, 36, 37, 38, 39^. In our study, we observed a significant novel association of SNP rs2973744, affecting *COL23A1* in 5% of ADEH+ patients. The SNP rs2973744 results in the presence of C instead of T at the splice donor site of an exon-intron boundary between exons 10 and 11 of the *COL23A1* transcript allele. SNPs at splice donor sites of several genes generally lead to exons skipping or generation of differentially spliced transcripts (Yamaguchi et al. 2002; Anna and Monika 2018). Interestingly, a SNP which leads to C instead of G at the splice donor site in thrombopoietin gene (*THRO*), resulted in elevated thrombopoietin protein (TPO) serum levels in patients with hereditary thrombocythaemia. This was an effect of more efficient translation of the resulting mutated transcripts than the wild type transcripts ^40^. Here we show that the heterozygous SNP in *COL23A1* results in a novel, differentially processed form of Col23a1 and, in the presence of other risk factors for EH, leads to higher transcript and total protein levels.

AD is a complicated multifactorial disease, resulting from genetic aberrations, abnormalities in the immunological response as well as environmental factors ^5, 6, 41^. A sub-group of patients suffering from AD is more prone to severe HSV-1 infections resulting in EH development ^7, 18, 20^. In line with that, we show here that primary keratinocytes isolated from an ADEH+ patient carrying the SNP rs2973744 were more susceptible to HSV-1 than those from ADEH- patients. Moreover, the *COL23A1* expression was increased in these patient-derived keratinocytes implying that higher levels of Col23a1 correspond to higher susceptibility to HSV-1.

Collagen type XXIII alpha 1 chain (Col23a1), encoded by *COL23A1*, is a type II transmembrane protein, with an N-terminal cytoplasmic domain, one hydrophobic transmembrane domain and three extracellular collagenous domains, flanked by short non-collagenous domains ^42^. It shares structural homology with collagen XIII and XXV. Col23a1 is a ligand for integrin α2β1 and is expressed in skin, brain, lung, cornea, tendon and kidney ^30, 43^. Alternative splicing results in three Col23a1 isoforms ^44, 45^ (Human Protein Atlas proteinatlas.org).

Col23a1 belongs to the class of non-fibrillar collagens and is located in its full-length form (75 kDa) in the lipid rafts of the cell membrane, or is cleaved by proteases, principally furin, in the Golgi/trans-Golgi network. The resulting ectodomain (60 kDa) is secreted from the cells into the extracellular matrix ^29, 42^. The shedding of Col23a1 depends on the tissue and the cholesterol content in the plasma membrane ^29^. Interestingly, the full-length Col23a1 is predominantly present in skin, whereas, in brain, the shed form of Col23a1 takes precedence, thus hinting at potentially divergent roles for the two forms ^43^.

Previous work showed that in cultured primary keratinocytes, Col23a1 was undetectable on the cell surface due to cleavage of the full-length protein by furin ^30^. Here, we also detected only low amounts of Col23a1 on the cell surface of primary foreskin keratinocytes. To study the effect of increased membrane levels of Col23a1 on HSV-1 infection, we reduced the shedding of Col23a1 by treating keratinocytes with FI and then infected them with HSV-1 in the absence of FI. Furin is a protease involved in the maturation and activation of envelope proteins of many viruses such as SARS-CoV-2, HIV, influenza, and Ebola. The proteolytic activation of viral proteins by furin can either occur during virus maturation, or viral entry into newly infected cells ^46^. Therefore, furin inhibitors are considered for their potential use as anti-viral drugs ^47^. In our study furin inhibition increased the susceptibility of keratinocytes to HSV-1, possibly by enhancing Col23a1 levels on the cell surface. Even though HSV-1 proteins do not contain furin cleavage sites, we cannot exclude that furin inhibition increased the susceptibility to HSV-1 by interfering with the proteolytic maturation of additional host factors ^46, 48^. As a second and more specific approach to study the effect of Col23a1 in HSV-1 infection, we overexpressed *COL23A1* in primary foreskin keratinocytes. Unfortunately, the cells stopped growing after transduction; therefore, we repeated the *COL23A1* overexpression in HaCaT cells with better success. Consistent with the FI experiment, we show here, that *COL23A1* overexpression significantly increased the susceptibility of HaCaT cells to HSV-1.

The clinical relevance for Col23a1 as a potential biomarker has been explored in several cancers ^49, 50, 51^. Knockdown of *COL23A1 in vitro* reduces cell adhesion and impairs anchorage-independent growth. Moreover, overexpression of *COL23A1* enhances cell adhesion in cancer cell lines, suggesting that Col23a1 promotes metastasis by aiding cell-cell and cell-matrix interactions and assisting anchorage-independent cell growth ^52^.

Interestingly, the class A macrophage scavenger receptor (MARCO), a protein distantly related to Col23a1, is reported to increase HSV-1 susceptibility in keratinocytes by enhancing virus binding to host cells ^53^. Moreover, MARCO-/- mice are protected against HSV-1 infections. Surprisingly, another study reported that epidermal sheets and primary dermal fibroblasts from MARCO-/- mice are as efficiently infected by HSV-1 as those isolated from control mice, suggesting that MARCO is not required for HSV-1 entry into the epidermis and dermal fibroblasts ^54^. However, to date, no studies have investigated the role of Col23a1 in viral infections. Therefore, our study is the first to report that elevated Col23a1 levels increase the susceptibility of keratinocytes to HSV-1.

To better understand the exact mechanism(s) behind increased HSV-1 gene expression in *COL23A1* overexpressing cells, we subjected the HaCaT cells to RNA sequencing. The transcriptome data revealed that *COL23A1* overexpression led to the downregulation of several genes, such as *IL1R1*, *IL32*, *TLR4*, *CFH*, *C3*, *S100A9*, *IRF1*, and *ADAM23* which are involved in the immune response of keratinocytes. Interestingly, upregulation of *TNC* (tenascin C) and *SPINK5* (serine peptidase inhibitor Kazal type 5), which are associated with AD ^33, 55^, was also observed in these cells. The mRNA levels of the above-mentioned genes were further validated using RT-qPCR.

In conclusion, our data show that elevated Col23a1 levels promote more efficient HSV-1 infection of keratinocytes presumably by attenuating their immune response. Consistently, AD patients carrying the heterozygous SNP rs2973744 are predisposed to develop severe HSV-1 infections. Early screening for the SNP rs2973744 in AD patients with recurrent HSV-1 infections might help to prevent disseminated HSV-1 infections through awareness and early therapeutic intervention.

## Methods

### Study subjects

The recruitment of patients suffering from AD with or without a history of EH was carried out at the Department of Dermatology and Allergy of Hannover Medical School, Germany. AD was diagnosed in the patients according to Hanifin and Rajka criteria ^56^. The ethics committee of Hannover Medical School (No. 8733_BO_S_2019) approved this study and all study participants provided written informed consent. DNA samples from AD patients with and without a history of EH were derived from the GENEVA cohort, Biobank of the University Hospital Schleswig-Holstein (UKSH), Kiel, Germany.

### Whole exome sequencing

Whole exome sequencing was performed at RCUG Genomics Core Facility of Hannover Medical School. The analysis involved 9 patients with a history of EH and first-degree healthy relatives of 7 patients. SureSelect^XT^ Target Enrichment System (Agilent, Santa Clara, CA, USA) and a NextSeq (Illumina, San Diego, CA, USA) sequencing device was used to perform sequencing. The analysis of data was performed using an established pipeline involving PrinSeq, FastQC, cutadapt, BWA, SAMtools or GATK. Genetic variants that occurred in more than one EH patient with a minor allele frequency <1% in the 1000 Genomes and ExAC databases and were rated ‘deleterious’, ‘possibly damaging’ or ‘probably damaging’ by MetaLR, Sift, PP2_HVAR, or PP2_HDIV, respectively were considered as the candidates. TaqMan-PCR was employed to re-analyze candidate variants in an age/sex-adjusted manner on three larger cohorts, comprising 117 AD patients with EH history, 117 AD patients who never experienced EH and 118 healthy controls from the POPGEN cohort and analyzed by ꭓ^2^ test (GraphPad Prism V5, Graphpad, LaJolla, CA, USA).

### Cell culture

Primary hair keratinocytes were derived from the outer root sheath of the hair follicle as previously described (Wang et al. 2008). The primary foreskin keratinocytes were isolated from the foreskin of anonymous children undergoing surgery as described (Wittmann et al. 2005). The consent of patients for experiments involving primary foreskin keratinocytes was not required as German laws consider leftover human tissue after surgery as discarded material. Primary hair and foreskin keratinocytes were cultured in Keratinocyte Growth Medium 2 (PromoCell, Heidelberg, Germany) supplemented with Keratinocyte Supplement Pack (KGM2; PromoCell). Primary hair keratinocytes were pre-stimulated with 1.4 mM CaCl_2_ (Carl Roth, Karlsruhe, Germany) for 24 h for each experimental set-up. MaxGel^™^ ECM (Merck, Darmstadt, Germany) was coated onto culture flasks to support the growth of primary hair keratinocytes. HaCaT cells, a spontaneously immortalized keratinocyte cell line ^57^, were cultivated in RPMI 1640 medium (Gibco by life technologies, Thermo Fisher Scientific, MA, USA) supplemented with 2 mM L-glutamine (Bio & Sell GmbH, Feucht, Germany), 100 µg/ml penicillin-streptomycin (Bio & Sell), 1x non-essential amino acids (Bio & Sell), 0.05 mg/ml gentamicin (Bio & Sell) and 6% (vol/vol) fetal bovine serum (FBS; PAN Biotech, Aidenbach, Germany).

BHK-21 [American tissue culture collection (ATCC), certified cell line 10 (CCL-10)] were cultured in minimum essential medium (MEM; Cytogen, Sinn, Germany) supplemented with 10% (vol/vol) feta calf serum (FCS; Life Technologies, Gibco, Darmstadt, Germany) and Vero cells (ATCC, CCL-81) were cultured in MEM with 7.5% (vol/vol) FCS.

### Viruses

We used either parental HSV1(17^+^)Lox (HSV-1) ^58^, HSV1(17^+^)Lox-_pMCMV_GFP (HSV1-GFP for short) ^59^, or HSV1(17^+^)Lox-_pMCMV_mChe (HSV1-Che for short) ^60^ expressing soluble GFP or mCherry under the control of the murine cytomegalovirus major immediate-early promoter, respectively. Virus was amplified by infecting BHK-21 cells with 0.01 pfu/cell. 60-90 hpi, virus was harvested from the supernatant of infected BHK, sedimented and titrated on Vero cells as described ^61, 62^.

### HSV-1 infection

To reach 80% confluency of cells on the day of infection, keratinocytes were plated one day before infection. Next day, cells were washed with PBS (PAN Biotech, Aidenbach, Germany) and KGM2 containing HSV-1 inoculum was added onto cells for 1 h and incubated at 37°C and 5% CO_2_. After 1 h incubation, the medium containing the virus was replaced with KGM2. A further incubation of 20 h was carried out at 37°C and 5% CO_2_. The virus strain and MOI used were assay-dependent and are described in the respective sections. HSV-1 infection was performed as described unless stated otherwise.

### mRNA isolation and RT-qPCR

Keratinocytes were lysed and RNA was isolated using the Reliaprep^™^ RNA Miniprep Systems (Promega, Germany) according to the manufacturer’s instructions. cDNA was prepared by reverse transcription of RNA using the RevertAid RT reverse transcription kit (ThermoScientific™, Dreieich, Germany). LightCycler^®^ 480 SYBR Green I Master (Roche Molecular Biochemicals, Manheim, Germany) was used to perform quantitative real-time PCR (qPCR) on a LightCycler 480 (Roche Molecular Biochemicals). Expression of *COL23A1, UL19, UL27, UL23, UL42, UL54, US12, IL1R1, IL32, C3,* and *TLR4* were quantified relative to the expression of the host gene *RPS20* (ΔΔCT). Primers were purchased from Qiagen (QuantiTect Primers; Hilden, Germany) and TIB Molbiol (Berlin, Germany). Primer sequences are given in the supplementary table S1.

### Flow cytometry

Keratinocytes infected with HSV1-GFP or HSV1-Che were washed twice with PBS, treated with 0.02% EDTA (5 min at 37°C; PAN Biotech, Aidenbach, Germany) and detached using 0.05% trypsin / 0.02% EDTA (5 min at 37°C; PAN Biotech). Trypsin inhibitor (PAN Biotech) was used to stop trypsinization, cells were then washed twice with 0.1% EDTA and fixed with 4% paraformaldehyde for at least 30 min at 4°C. GFP and mCherry expression were analyzed by flow cytometry (CytoFLEX, Beckman Coulter, CA, USA).

To detect Col23a1 cell surface levels, keratinocytes were washed twice with PBS, treated with 0.02% EDTA for 5 min and detached using Accumax (20 min at 37°C; PAN Biotech). Cells were incubated with either 8 µg/ml IgG_2B_ isotype (cat. #MAB0041; R&D Systems, MN, USA) or mouse monoclonal IgG_2B_ antibody against human collagen XXIII α1 (cat. #MAB4165; R&D Systems, MN, USA) for 45 minutes at 4°C. Cells were then labeled with 1 µg/ml donkey polyclonal anti-mouse secondary IgG antibody (Alexa Fluor^®^ 647 (AF647) fluorochrome) (Abcam, Cambridge, UK) for at least 30 min at 4°C. AF647 fluorescence was measured by flow cytometry (FACSCalibur™, Becton Dickenson, NJ, USA).

To measure intracellular Col23a1 levels, keratinocytes were trypsinized as mentioned before. The protocol for the eBioscience™ Foxp 3/ Transcription Factor Staining Buffer Set (Invitrogen™, Thermo Fisher Scientific, MA, USA) was followed to stain intracellular Col23a1 levels by flow cytometry (CytoFLEX™, Beckman Coulter, CA, USA) with the same antibodies used for detecting Col23a1 cell surface levels.

### Protein lysate preparation and Western blotting

Cells were lysed on ice with M-PER^™^ Mammalian Protein Extraction Reagent (Thermo Scientific™) containing protease inhibitor Pefabloc^®^ SC (Merck) and incubated on a shaker for 5 min at 4°C. Cells were scraped off the culture plate and the contents were subjected to centrifugation at 14,000 x *g* for 10 min at 4°C. Supernatants were collected and protein concentrations were estimated using Qubit^™^ Protein Assay kit (Invitrogen, MA, USA). Equal amounts of protein were loaded on 4-20% SDS-polyacrylamide gel (Bio-Rad Laboratories, CL, USA). The protein bands were transferred to the polyvinylidene fluoride membrane and blocked for 1 hour at room temperature with 5% milk. Proteins were probed using monoclonal mouse anti-GAPDH antibody (cat. #14C10; Cell Signaling Technologies, MA, USA) or monoclonal mouse IgG_2B_ anti-human collagen XXIII α1 antibody (cat. #MAB4165; R&D Systems) and labeled with secondary anti-mouse IgG HRP-linked antibody (cat. #7076; Cell Signaling Technology). The blots were developed with SignalFire^™^ Elite ECL Reagent (Cell Signaling Technology) and images were captured using ChemiDoc Imaging System (Bio-Rad Laboratories). For quantification, the Image Lab software (Bio-Rad Laboratories) was used.

### Furin inhibitor I

Primary foreskin keratinocytes were seeded to reach 80% confluency on the day of treatment. 75 µM of Furin inhibitor I (FI) – Decanoyl-Arg-Val-Lys-Arg- chloromethylketon (Dec-RVKR-CMK; Bachem, Bubendorf, Switzerland) was added to the cells and incubated for 24 h at 37°C and 5% CO_2_. The next day, cells were either used to analyze Col23a1 surface expression or their susceptibility to HSV1-GFP (0.75 pfu/cell) using flow cytometry.

### Overexpression of *COL23A1*

Control lentiviral particles (#LPP-NEG-Lv205-100) encoding EGFP and a puromycin resistance cassette and lentiviral particles encoding *COL23A1* (#LPP- Y2210-Lv205-100) in addition were purchased from GeneCopoeia, Rockville, MD, USA. One day before transduction, HaCaT cells were seeded to reach 40% confluency on the day of transduction. Control and *COL23A1* lentiviral particles were incubated in transduction media containing RPMI and 3 µg/ml polybrene (Merck) for 20 min at room temperature. Then, Y- 27632 dihydrochloride (Rho kinase inhibitor; Abcam, Cambridge, UK) was added to the lentiviral particles at a final concentration of 10 µM. This inoculum was transferred onto HaCaT cells (24 transducing units/cell) and incubated overnight at 37°C with 5% CO_2_-atmosphere. The following day, media was replaced with fresh RPMI containing 10 µM Rho kinase inhibitor and cells were incubated further at 37°C and 5% CO_2_. 3 days after transduction, 15 µg/ml puromycin was used for the selection of transduced cells. Overexpression was evaluated using RT-qPCR, flow cytometry and Western blot.

### HSV-1 cell-to-cell spread analysis

HaCaT cells were seeded to reach 100% confluency on the day of infection. Cells were infected with HSV1-Che at very low MOIs of 0.001, 0.01 and 0.05 pfu/cell. At 20 hpi, cells were washed and fixed with 4% paraformaldehyde. Infection centers were visualized using the BioTek Cytation 5 Cell Imaging Multimode Reader, and plaque size and Cherry intensity were quantified using CellProfiler software (Carpenter et al. 2006).

### HSV-1 temporal gene expression analysis

HaCaT cells were seeded to reach 100% confluency on the day of infection. Cells were infected with 5 pfu/cell of HSV-1 for 1 h at 37°C and 5% CO_2_. After 1 h, media containing virus was replaced with fresh KGM2 and incubated further for 2 h (data not shown) 4 h, 6 h and 8 h. After 2, 4, 6 and 8 hpi, cells were lysed, RNA was isolated, and the expression of HSV-1 genes relative to the host gene *RPS20* was measured by RT-qPCR.

### Bulk RNA-seq data analysis

Control and *COL23A1* overexpressing HaCaT cells were infected with 5 pfu/cell of HSV-1. At 2 hpi and 6 hpi, RNA and cDNA was prepared from cells as mentioned before. cDNA libraries were generated using ‘NEBNext^®^ Ultra II Directional RNA Library Prep Kit for Illumina’ (E7760L; New England Biolabs) and sequenced using Illumina NextSeq 550 sequencer. The raw data files were processed using nfcore/rnaseq (version 3.9) and expression analysis was performed on the internal Galaxy (version 20.05). R package DESeq2 (version 1.42.1) was used to identify differentially expressed genes from raw gene counts, running on R version 4.3.3 within the RStudio IDE (version 2023.12.1+402) ^63^. A single-factor design was used in DESeq2. EnhancedVolcano package (version 1.20.0) was used to generate volcano plots of differentially expressed genes with a log2(FC) value of >0.5 or <- 0.5 and heatmaps of the normalized counts of selected genes. Gene set enrichment analysis on our list of differentially expressed genes to identify significantly enriched KEGG and GO biological processes was performed using R package ClusterProfiler (version 4.10.1) ^64^.

### Statistical analysis

Data was assessed for normality and subsequently two-tailed Paired t-test (parametric test) or Mann-Whitney *U* test were used to compare group means or medians. Statistical analyzes and plots were generated using GraphPad Prism (version 8.0, GraphPad Software, San Diego, CA). Asterisks in the figures indicate p-values: \**P* < 0.05, \*\**P* < 0.01, \*\*\**P* < 0.001. Box & whiskers plot represent the 10-90 percentiles.

## Data Availability

All data produced in the present study are available upon reasonable request to the authors.

## Abbreviations

AD: atopic dermatitis
ADEH+: AD patients with a history of eczema herpeticum
ADEH-: AD patients without a history of eczema herpeticum
Che: mCherry
Col23a1: collagen type XXIII alpha 1 chain
E: early HSV-1 gene
EH: eczema herpeticum
FI: furin inhibitor I
GFP: green fluorescent protein
GO: Gene Ontology
Hpi: hours post-infection
HSV-1: herpes simplex virus type 1
IE: immediate-early HSV-1 gene
L: late HSV-1 gene
MARCO: macrophage receptor with collagenous structure
MOI: multiplicity of infection
mRNA: messenger RNA
PCR: polymerase chain reaction
Pfu: plaque-forming unit
qPCR: quantitative PCR
RNA-seq: RNA sequencing
RT-PCR: reverse transcription PCR
RT-qPCR: quantitative RT-PCR
SD: standard deviation
SNP: single nucleotide polymorphism
WT: wild type.

## Acknowledgements

Shruti Chopra was supported by the Hannover Biomedical Research School (HBRS) and the Center for Infection Biology (ZIB). This study was funded by EXNet-0030-Phase2-3 Communities allied in infection (COALITION) and by the Deutsche Forschungsgemeinschaft (DFG, German Research Foundation) under Germany’s Excellence Strategy-EXC 2155 “RESIST”- project ID 39087428. Sequencing data used in this publication was generated by the Research Core Unit Genomics (RCUG) at Hannover Medical School, and we especially thank Lutz Wiehlmann and Oliver Dittrich-Breiholz. We express our gratitude to Winfried Hofmann, Gunnar Schmidt, Doris Steinemann and Lina Werfel from Hannover Medical School for their invaluable advice and insightful discussions, and to Petra Kienlin, Gabriele Begemann, and Ilona Klug for their excellent technical support.

## Author Contributions

J.Z., K.D., L.M.R., S.C., T.F.S., and T.W. designed the experiments. S.C. performed the experiments. J.Z., K.D., L.M.R., and S.C. carried out the analysis and interpretation of the data. E.R., I.H., and L.M.R. performed genomics data analyses. S.W. and W.L. provided samples. B.S. and K.D. provided resources. S.C. and S.T. performed the transcriptomics data analyses. J.Z., K.D., L.M.R., and T.W. supervised the project. S.C. wrote the first draft of the manuscript. All authors reviewed and edited the manuscript.

