## Supplementary Table S1 and Figure S1 for "Identification and characterization of collagen XXIII alpha 1 as a novel risk factor for eczema herpeticum"

**Table S1.** Primers used for the detection of transcripts.

| Target | Forward (5´-3´) | Reverse (5´-3´) |
|---|---|---|
| Human *RPS20* | QuantiTect Primers (Qiagen, Hilden, Germany) | |
| Human *COL23A1* | QuantiTect Primers (Qiagen, Hilden, Germany) | |
| Human *IL1R1* | QuantiTect Primers (Qiagen, Hilden, Germany) | |
| Human *IL32* | QuantiTect Primers (Qiagen, Hilden, Germany) | |
| Human *TLR4* | QuantiTect Primers (Qiagen, Hilden, Germany) | |
| Human *CFH* | QuantiTect Primers (Qiagen, Hilden, Germany) | |
| Human *C3* | QuantiTect Primers (Qiagen, Hilden, Germany) | |
| Human *S100A9* | QuantiTect Primers (Qiagen, Hilden, Germany) | |
| Human *IRF1* | QuantiTect Primers (Qiagen, Hilden, Germany) | |
| Human *ADAM23* | QuantiTect Primers (Qiagen, Hilden, Germany) | |
| Human *TNC* | QuantiTect Primers (Qiagen, Hilden, Germany) | |
| Human *SPINK5* | QuantiTect Primers (Qiagen, Hilden, Germany) | |
| HSV-1 *US12* | GTCCTCACGCCCCCTTTTAT | GAAATGGCGGACACCTTCCT |
| HSV-1 *UL19* | TCTCAGTCACAAAGCGGTCC | TTCAAGATCAGTCCCGTGGC |
| HSV-1 *UL23* | GGTCATGCTGCCCATAAGGT | CTCACCCTCATCTTCGACCG |
| HSV-1 *UL27* | CACAGGGTCAGCTCGTGATT | CATACAGCGCCATGTCAACG |
| HSV-1 *UL42* | ACTCGCTTCTGGTTATGGGC | ACCGACTGAATTGCGAGTGT |
| HSV-1 *UL54* | CTCCAGTGCTACCTGAAGGC | TCCTTAATGTCCGCCAGACG |

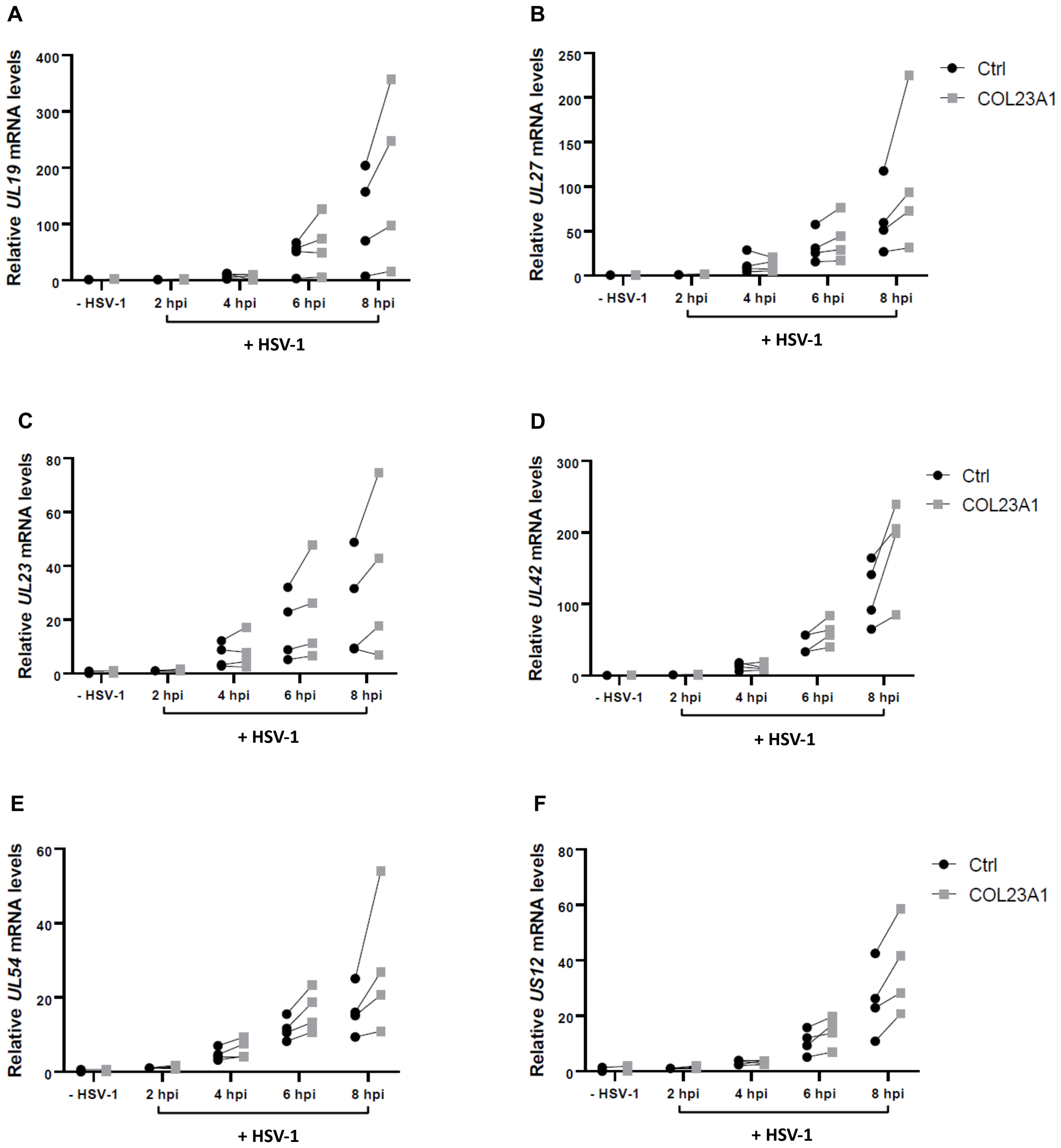


**Figure S1: *COL23A1* overexpression enhances HSV-1 transcript levels.** (A-F) Control and *COL23A1* overexpressing HaCaT cells were infected with 5 pfu/cell of HSV-1. *UL19, UL27, UL23, UL42, UL54 and US12* mRNA levels were measured at 2 hpi, 4 hpi, 6 hpi, and 8 hpi by RT-qPCR and normalized to the housekeeping gene *RPS20* (black bars, mean ± standard deviation [SD]; n = 4 independently repeated experiments). Ctrl= HaCaT cells stably transduced with control lentiviral particles; COL23A1= HaCaT cells stably transduced with *COL23A1* lentiviral particles.
